# Effect of a gender-synchronized family planning intervention on inequitable gender norms in a cluster randomized control trial among husbands of married adolescent girls in Dosso, Niger

**DOI:** 10.1101/2023.09.28.23296292

**Authors:** Sabrina C. Boyce, Alexandra M. Minnis, Julianna Deardorff, Sandra I. McCoy, Sneha Challa, Nicole E. Johns, Sani Aliou, Mohamad I. Brooks, Abdoul-Moumouni Nouhou, Holly Baker, Jay G. Silverman

## Abstract

Gender inequity is pervasive globally and has severe consequences for health and well-being, particularly for women and girls in Niger. The Reaching Married Adolescents in Niger (RMA) intervention aimed to promote equitable gender norms in order to increase modern contraceptive use and reduce intimate partner violence among married adolescent girls and their husbands in Niger. Using data from a 4-arm factorial cluster randomized control trial of the RMA intervention (2016-2019), the current study assesses effects of the RMA intervention on gender norms among husbands. We used an adjusted hierarchical difference-in-differences linear regression model to assess these effects. The mean score for perceived gender inequitable norms at baseline was 4.1 (n=1,055; range: 0-5). Assignment to the RMA small groups intervention was associated with a 0.62 lower score (95% CI: −1.05, −0.18) relative to controls at follow-up, after adjusting for baseline differences. No significant effects were detected for other intervention arms. As a low-cost, simple, scalable, and transferrable intervention with rigorous evidence of being able to change such gender norms, this community health worker-based small group intervention could be valuable to the field of public health for reducing the negative impact of inequitable gender norms on health and wellbeing in similar settings.

## 1. INTRODUCTION

Gender inequity is pervasive globally and has severe consequences for health and well-being (Heise, 2019). The relationship between gender inequity and related poor health may be especially clear in Niger, which ranks 154^th^ of 162 countries on the United Nations Gender Inequality Index, based on reproductive health, empowerment, and economic activity (UNDP, 2020). With 76% of Nigerien girls marrying by the age of 18 and an adolescent birth rate of 177 births per 1000 adolescents aged 15-19, early and frequent childbearing has contributed to an overall birth rate of 6.7 births per woman and maternal mortality ratio of 509 (per 100,000 live births) (The World Bank, 2020; UNICEF, 2018; UNPD, 2019; WHO, 2019). Early marriage and childbearing create family power structures in which married adolescent girls have minimal decision-making power relative to their older husbands and mothers-in-law and relatedly, limited ability to control their reproductive and sexual health, including the decision to use modern contraceptive methods (Fleming et al., 2020; Girls Not Brides, 2015; Tomar et al., 2021; UNICEF, 2014). Gender role expectations in this context confine married adolescent girls to the household, to care for the home and bear children early and often, and away from educational and leadership opportunities (Perlman et al., 2018). In Niger, as well as globally, shifts toward gender equity are viewed as crucial for societal well-being (Heise, 2019).

Gender inequity is embodied in the social norms of a society, which are expressed across all levels of Brofenbrenner’s socio-ecological framework (individual, social, institutional, and macro) (Bronfenbrenner, 2005; Cislaghi & Heise, 2019). Understood through social norms theory, social norms are the unspoken rules in a society about what behaviour is acceptable and unacceptable (Mackie et al., 2015). In highly patriarchal contexts, gender norms, a subset of social norms that define gender-based social roles and behaviour, often condone dominant forms of masculinity, female submissiveness, and male entitlement to control of women’s bodies, even when these norms do not yield the best outcomes for the people involved (Bussey & Bandura, 1999; Cislaghi & Heise, 2017; Keleher & Franklin, 2008). The centrality of social norms, particularly gender norms, in shaping gender inequity and related practices is emphasized in multiple conceptual frameworks that inform behavioural interventions (Cislaghi & Heise, 2019; Heise, 1998; Pulerwitz et al., 2019). Gender norms and gender-based power simultaneously shape and are shaped by the institutions, resources, and social and individual systems of a society (Cislaghi & Heise, 2019).

Evidence suggests that public health interventions that change social norms, alongside other elements in the socioecology, can reduce the harmful health effects of gender inequity, but major gaps exist in understanding what intervention approaches and programs work to change social norms (Cislaghi & Heise, 2019; Dworkin et al., 2013; Haylock et al., 2016; Heise, 2019). There is growing interest in approaches that target social norms to reduce the health consequences associated with gender inequity (e.g., gender-based violence, child marriage, female genital cutting), especially in low- and middle-income countries (Cislaghi & Heise, 2019). For example, social norm interventions targeting both men and women, like Gender Equity Movement in Schools in India, Stepping Stones in South Africa, and Tostan in Senegal, have been shown to reduce intimate partner violence (IPV), herpes simplex 2 (HSV-2) infection, and female genital cutting, among other important health outcomes (Achyut et al., 2015; Jewkes et al., 2008; Rowley E, 2020). Increasing interest in the involvement of men, often the decision-makers, gate-keepers, and perpetrators of violence against women, in gender norm-focused interventions is motivated by the potential of their involvement to disrupt and transform traditional power imbalances between men and women (Dworkin et al., 2013; Gupta, 2000). Synthesizing many of these learnings, the Social Norms Learning Collaborative (IRH, 2017) has proposed ten key attributes for social norm-shifting interventions, including creating safe space for critical community reflection, addressing power imbalances and inequity, and emphasizes the creation of a new positive norm. Little rigorous evidence exists, however, of social norms-focused interventions that have actually changed the targeted social norms, even when the primary health outcomes are observed to improve.

Measurement of social norms is difficult and contributes to why program impact on social norms is rarely assessed (Cislaghi & Heise, 2016; Mackie et al., 2015). Cialdini et al.’s pivotal work around social norms has led to the classification of social norms as either *descriptive* (i.e., beliefs about what others do), *injunctive* (i.e., beliefs about how others would respond to one’s behaviour), or *second order beliefs* (i.e., beliefs about what others approve and disapprove of), all of which require careful consideration for accurate measurement (Chung, 2016; Cialdini et al., 1991; Cialdini, 1998; Mackie et al., 2015). In the rare cases in which effects on social norms are considered, aggregates of individual attitudes are often used as a proxy measure of social norms, which fails to capture the core element of social norms: perceived social expectations around behaviour (Cislaghi & Heise, 2016). The evaluation of SASA!, a community mobilization intervention to prevent violence and reduce HIV-risk behaviours first implemented in Uganda, is justifiably acclaimed as one of few cluster randomized control trials (cRCT) to assess and observe program effects on both social norms and behavioural outcomes, yet measurement of community-level social norms relied on aggregates of individual attitudes (Abramsky et al., 2014). The quasi-experimental evaluation of Voices of Change in Nigeria, a program that targeted young women and men to strengthen enabling environments for gender equality, assessed program effects on social norms using measures that include social acceptability of behaviour, but effects were not detected (Denny et al., 2017). Similarly, The Saleema Initiative in Sudan, which aimed to promote community-level abandonment of female genital cutting, and Communities Care Programme in Somalia, which aimed to change social norms to end violence against women and girls in conflict-affected communities, showed quasi-experimental evidence of changing gender norms, with robust measurement of norms rather than aggregated attitudes, yet effects on the intended behaviours (female genital cutting and IPV, respectively) were not detected (Evans et al., 2019; Glass et al., 2019). As momentum to use social norm approaches in LMICs grows (Cislaghi & Heise, 2019), it is critical for researchers and practitioners to understand whether social norms-focused interventions actually change their intended social norms, and if so, whether behaviours informed by these norms subsequently change as a result. To advance this burgeoning field in public health research, such evidence is critical to supporting the development of programmatic conceptual frameworks and of social norm-shifting interventions.

The Reaching Married Adolescents in Niger (RMA) intervention, implemented starting in April 2017, aimed to address social norms and other barriers to increasing modern contraceptive use and reducing IPV among married adolescent girls and their husbands in the Dosso region of rural Niger. These primary behavioural outcomes were measured after receipt of one year of program implementation and evidence of positive effects on both outcomes and diffusion of the contraceptive use effects among social network members have been reported elsewhere (Shakya et al., 2021; Silverman et al., 2022). The current study assesses intervention effects on gender norms among husbands, the primary decision-makers around contraceptive use and perpetrators of IPV. Findings will help to close the gap in evidence on what works to change gender norms in Niger and similar contexts.

## 2. MATERIALS AND METHODS

### 2.1. Intervention

The RMA intervention is a gender-synchronized (i.e., concurrent programming for men and women), community-based program, developed and implemented by Pathfinder International among married adolescent girls (ages 13-19) and their husbands in the Dosso region of Niger to increase healthy birth spacing via use of modern contraceptive methods. Program approaches, which were randomly assigned, were household visits or small group discussions; community-level dialogues were held concurrently in all intervention arms. Household visits were conducted by gender-matched, trained community health workers (CHWs) via 12 monthly visits to individual households for both married adolescent girls and their husbands. Household visits, provided to wives and their husbands separately, provided information and counselling on healthy timing and spacing of pregnancies and on access to and use of modern contraceptive methods. Small group discussions, also held separately by gender, were conducted by gender-matched CHWs trained to facilitate small groups and were held twice monthly for wives and once monthly for their husbands. Content delivered in these groups included general health and life skills, reproduction anatomy and health, use of modern contraceptive methods to accomplish healthy timing and spacing of pregnancies, gender norms that impede contraceptive use and female autonomy, couples’ communication regarding fertility decisions, and gender-based violence. Community dialogues were convened at the village-level each month by two trained facilitators to engage community gatekeepers and key influencers (e.g., traditional and community leaders, parents and in-laws) in creating an environment supportive of healthy timing and spacing of pregnancies and contraceptive use among married adolescent girls and their husbands (more details on the intervention, sample, and trial design are available elsewhere) (Challa et al., 2019). The intervention was implemented for 12 months.

### 2.2. Research Study Design

We conducted a 4-arm factorial cRCT to evaluate the impact of receiving one year of the RMA intervention in 48 villages in the Loga, Doutchi, and Dosso districts in the Dosso region of Niger. A two-staged, stratified randomization strategy was utilized to accommodate the districts approved by the Niger Ministry of Health for inclusion and the limited feasibility of implementing more than one intervention model within a district. In the first randomization step, each of the three districts were randomly assigned a treatment condition (1:1:1). Arm 1 received household visits (Loga), Arm 2 received small group discussions (Doutchi), Arm 3 received both (Dosso). All intervention arms also received community dialogues.

In the second randomization step, randomly selected villages within each district were assigned to either the treatment assigned to that district or the control group. Within each district, 16 villages were randomly selected for inclusion from villages meeting inclusion criteria: 1) having at least 1000 permanent inhabitants; 2) primarily Hausa or Zarma-speaking (the two major languages of Niger); and 3) not receiving any intervention specific to contraceptive use or gender equity in the past two years. Within each district, four of the 16 randomly selected villages were randomly assigned to the control group and the remaining 12 to the intervention condition assigned to that district, such that 12 villages were assigned to each study arm (1:1:1:1). Arm 4 therefore consisted of the four control villages in each district (12 villages total). Random allocation was done by US-based study staff, with limited familiarity of the region at the time, by assigning numbers sequentially to an alphabetical list of the 16 villages and using a computer-generated random selection of four of those numbers for assignment to the control arm. The chief in each village provided a complete list of all households in which a married adolescent aged 13-19 years lived and from this list, 25 wife-husband dyads were randomly selected for recruitment to the study, using a similar computer-generated random number selection process by US-based study staff. Local, trained research assistants visited the selected households to confirm eligibility (being Hausa or Zarma speaking; not planning to move away from the village in the next 18 months; not planning to travel away from the village for more than 3 months during that period; not sterilized; and providing informed consent) and random replacement was used if a household was not eligible or available.

The number of villages and wife-husband dyads selected for inclusion in the study was based on a power calculation for a minimum effect size on the main outcome, modern contraceptive use, of 2.0 or greater odds associated with the intervention. This calculation accounted for four arms with 12 clusters of equal size nested within each arm, time-by-treatment effects, an intra-class correlation kappa value of 0.05, attrition of 10%, and 80% power to detect an association of this power (Challa et al., 2019).

Data utilized in this analysis were collected at baseline (T1; May-June 2016) and 24-month follow-up (T2; April-June 2018). Full implementation of the intervention began 12 months after baseline data were collected and T2 was conducted after 12 months of exposure to the intervention. At both time points, local research assistants who spoke Hausa or Zarma (depending on the village’s language) conducted data collection using questionnaires programmed on tablet computers. Gender-matched research assistants interviewed married adolescent girls and husbands separately in a location deemed audibly private. The survey took 45-60 minutes to complete.

This trial was pre-registered on clinicaltrials.gov (NCT03226730) on July 24, 2017, with social norms regarding gender inequity defined as a secondary outcome. Ethics review boards of the University of California, San Diego School of Medicine (Project 160407S) and the Niger Ministry of Health (Project 011/2016/CCNE) approved all study protocols.

### 2.3. Sample

There were 48 villages in the sample, with an average of 21 wife-husband dyads per village (range: 18 - 27 dyads). Only data provided by husbands were included in this analysis as only participants who reported a male gender identity were asked the evaluated items on social norms regarding gender inequity. Data from female-identifying participants regarding RMA effects have been previously reported (Silverman et al., 2022).

### 2.4 Measures

Perceived social norms regarding gender inequity (henceforward, SN-GEMS) was measured using a 5-item scale of second-order beliefs, a type of social norm, among husbands (Cronbach’s alpha at T1 and T2: 0.62). The social norms measure was created for this study, adapted from the Gender Equitable Men Scale (GEMS), which has been used in more than 20 countries to assess male attitudes regarding gender equity (Singh et al., 2013). We adapted this scale by adding a stem to each dichotomous item that read, “People in my community think that…,” in order to measure second order beliefs. The items then proceeded as, 1) “…a woman’s most important role is to take care of the home and cook for the family”; 2) “…a man should have the final word about decisions in the home”; 3) “…there are times when a woman deserves to be beaten”; 4) “…a woman should never question her husband’s decisions even if she disagrees with them”; 5) “…it is natural and right that men have more power than women in the family.” Response options included “agree”, “disagree”, “don’t know”, and “decline to answer”. Responses of “don’t know” and “decline to answer” were treated as missing for that item. Scores were calculated by adding up the number of affirmative responses a participant gave to the five items, with missing items assigned a zero value (range: 0-5); a higher score indicates higher perceived social acceptability of inequitable gender norms.

Covariates were included in adjusted models to control for baseline socio-demographic differences between intervention and control arms. Factors considered were those that were identified to be associated with at least one intervention arm, via unadjusted multinomial logistic regressions for each factor (p<0.05). Those that were selected were hypothesized, based on content expertise and theory, to also be related to the outcome. Covariates included husband’s age, husband’s and wife’s education levels, husband’s migration status, total household assets, and district (because intervention assignment was randomized at the district level). In calculating inverse probability of censoring weights (IPCW), used to minimize possible selection bias due to loss to follow-up, we also included covariates representing baseline differences between husbands retained and those lost to follow-up at T2. These covariates included husband’s age (continuous), husband’s age at marriage (continuous), husband’s parity (continuous), husband’s and wife’s education level, and husband’s migration status. *Husband’s* and *wife’s education levels* were categorically measured as ever receiving any modern/government school, any Quranic school (Muslim religious education), or no schooling (reference category). *Total household assets,* a measure of wealth, was measured as a count of how many of six listed items (e.g., mobile phone, bike, animal-drawn cart) any member of the household owned and included as a continuous variable (range: 0-6). *Husband’s migration status* was dichotomously assessed as whether the husband spent more than 3 months away from the village in the past 12 months.

### 2.5. Data Analysis

To account for the two-step randomization process, imbalances at baseline, and loss-to-follow-up, we assessed the intention-to-treat effect of the RMA intervention on husbands’ SN-GEMS score using a hierarchical difference-in-difference linear regression. The primary model adjusted for covariates, accounted for village-level clustering in the study design and for multiple observations, and used IPCW to account for loss to follow-up. The model used robust standard errors to account for possible misspecification of the covariance structure.

Inverse probability of censoring weights were constructed by taking the inverse of the predicted probability of being retained in the study based on baseline demographic characteristics (Robins et al., 1995). Predicted probabilities were estimated using a logistic regression model that regressed study retention on covariates associated with loss to follow-up (covariates indicated for weights above). Missing weights were replaced with median weight value. In the model, these weights rebalance the sample such that follow-up participants who had low predicted probabilities of being retained in the study are up-weighted and those participants who had high predicted probabilities are down-weighted.

All analyses used STATA 15.1 (StataCorp, 2017). Significance was set at p<0.05 for statistical testing and estimates with 95% confidence intervals (CIs) are reported throughout.

## 3. THEORY

The theoretical basis for the Reaching Married Adolescents intervention is Fishbein and Yzer’s modified Theory of Reasoned Action that posits how shifts in behavioural, normative, and efficacy beliefs influence one’s attitudes, perceived social norms, and self-efficacy (Challa et al., 2019; Fishbein & Yzer, 2003). Together, the attitudes, perceived norms, and self-efficacy inform behavioural intentions, and combined with their skills and environmental constraints, these shifts in behavioural intentions lead to shifts in behaviour. Through the educational sessions of the RMA small groups and household visits, CHWs promote family planning and gender equity within families, which aims to shift behavioural, normative, and efficacy beliefs toward behavioural intentions that include contraceptive use and non-violent interpersonal relationships. This research assesses whether the RMA intervention succeeded in shifting perceived social norms, a critical step toward achieving the desired behaviour change.

## 4. RESULTS

At baseline, 1080 of 1351 eligible husbands completed surveys (79.9% male participation) and 773 participated in data collection at T2 (71.6% male retention) (Figure 1. CONSORT Participant Flow) (Silverman et al., 2022). On average, husbands were 25.6 (SD=5.3) years old at baseline (Table 1). About one-third of husbands (29.4%) had never received any schooling. Husbands had 1.5 children on average and 67.2% migrated for more than three months in the past year. Households reported owning on average 2.1 (SD=1.2) of the listed 6 assets.

**Figure 1.**
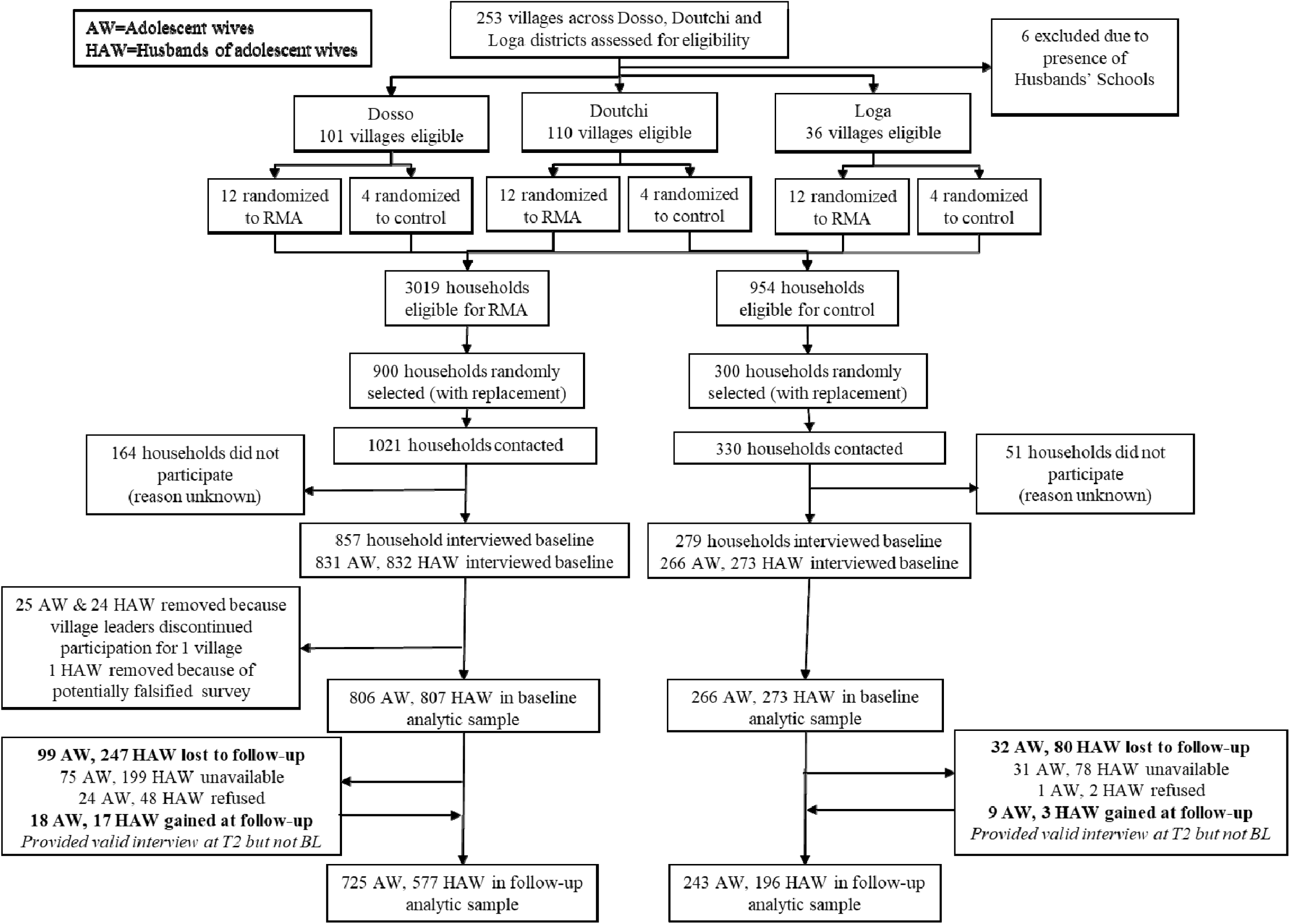
CONSORT Participant Flow for the Reaching Married Adolescents in Niger cluster randomized control trial (2016-2019)

**Table 1.** Husband participation in the Reaching Married Adolescents in Niger (RMA) Household Visits and Small Group Intervention Components at 24-month Follow-Up <u>(Dosso, Niger)</u>.

| **Husband RMA**<br>**Program Exposure** | **RMA Intervention Arm** |  |  | Pearson's<br>Chi-Square |
| --- | --- | --- | --- | --- |
|  | Arm 1: Household<br>visits<br>(n=205) | Arm 2: Small<br>groups<br>(n=194) | Arm 3:<br>Combination<br>(n=177) |  |
| Participation in at least<br>one RMA intervention<br>component | 146 (71.2%) | 125 (64.4%) | 134 (75.7%) | p=0.06 |
| 0 household visits | 59 (28.8%) | - | 69 (39%) |  |
| 1-3 household visits | 71 (34.6%) | - | 62 (35%) |  |
| 4+ household visits | 75 (36.6%) | - | 46 (26%) |  |
| 0 small groups | - | 69 (35.6%) | 60 (33.9%) |  |
| 1-3 small groups | - | 66 (34%) | 56 (31.6%) |  |
| 4+ small groups | - | 59 (30.4%) | 61 (34.5%) |  |

### 4.1. Participation

Participation in 12 household visits and/or small groups (one per month for one year) represents full receipt of the possible RMA intervention. In Arm 1, 71.2% of husbands participated in at least one program element, 64.4% in Arm 2, and 75.7% in Arm 3 (Table 1). A Pearson’s Chi Square test indicated a non-significant difference between participation in at least one program element across intervention arms, with the lowest participation among husbands in Arm 2 (p=0.06). About one-third (29.7%) of the sample assigned to an intervention did not participate in any component of the evaluated intervention.

### 4.2. Baseline differences across study arms

In comparing each intervention arm to the control arm, demographic characteristics varied (Table 2). Levels of education among husbands and wives were significantly different in all arms relative to controls. Arm 1 had lower Quranic education for men and women (p<0.001) relative to control, Arm 2 had higher government and Quranic education (p<0.001), and Arm 3 had lower Quranic education for women (p<0.05) relative to controls. Total assets were lower in Arms 1 and 3 (p<0.01 and p<0.05, respectively) while Arm 2 had lower levels of husband migration (p<0.001), relative to controls. Relative to controls, wives in Arm 2 were younger when married (13.8 years in Arm 2 vs. 14.3 years in control arm, p<0.01) and at baseline (17.0 years in Arm 2 vs. 17.4 years among controls, p<0.01) and husbands were older at baseline (26.5 years in Arm 2 vs. 25.5 years in control arm, p<0.05; *results not shown*). In terms of the outcome, perceived inequitable gender norms were substantially lower at baseline in Arm 1 compared with the control arm (p<0.001) (Table 2).

**Table 2.**
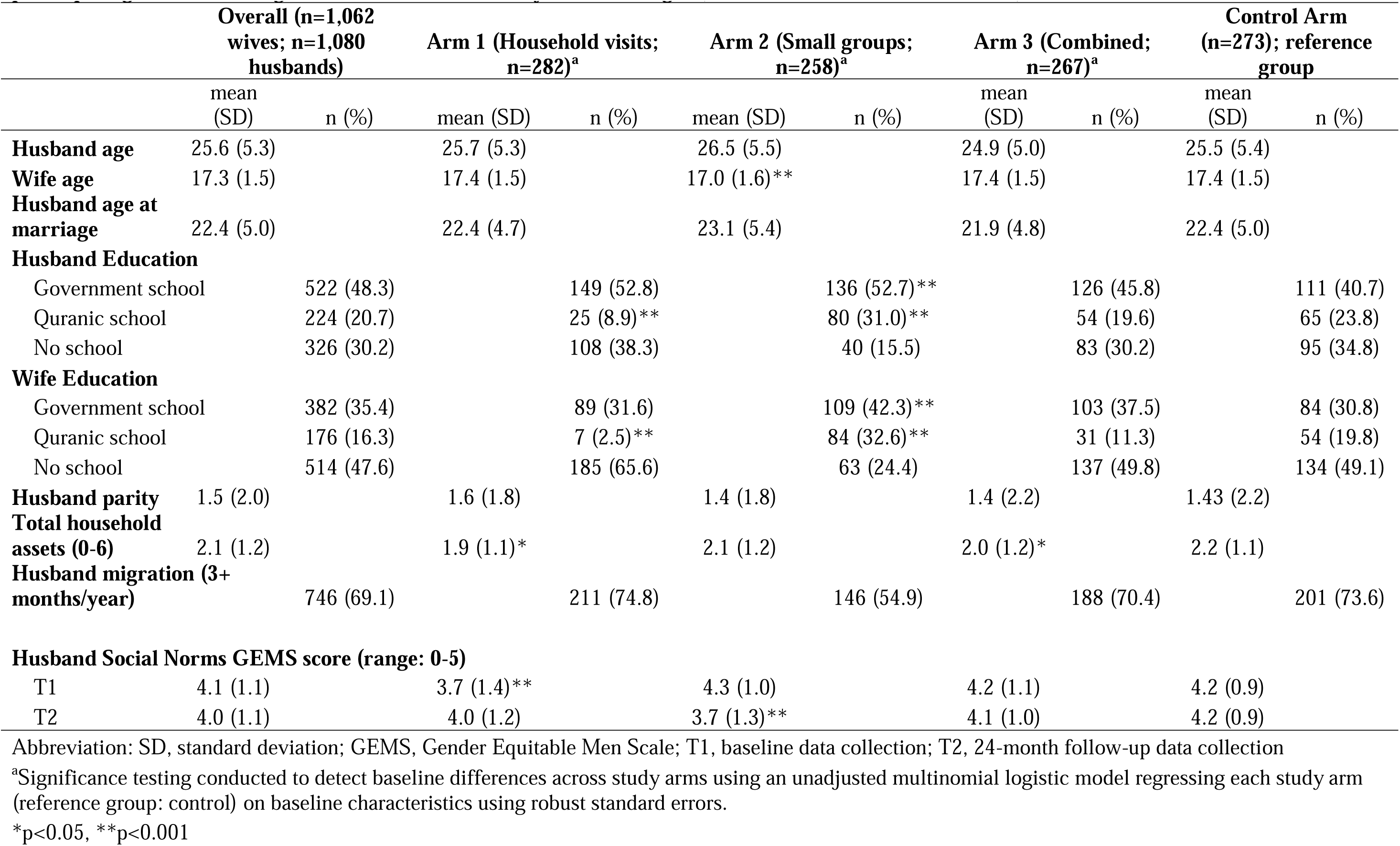
Baseline socio-demographic characteristics and perceived inequitable gender norms among married adolescents girls and their husbands participating in the Reaching Married Adolescents Study in Dosso, Niger (n=1,080 husbands; n=1,062 wives)

|  | Overall (n=1,062<br>wives; n=1,080<br>husbands) |  | Arm 1 (Household visits;<br>n=282)<sup>a</sup> |  | Arm 2 (Small groups;<br>n=258)<sup>a</sup> |  | Arm 3 (Combined;<br>n=267)<sup>a</sup> |  | Control Arm<br>(n=273); reference<br>group |  |
| --- | --- | --- | --- | --- | --- | --- | --- | --- | --- | --- |
|  | mean<br>(SD) | n (%) | mean (SD) | n (%) | mean (SD) | n (%) | mean<br>(SD) | n (%) | mean<br>(SD) | n (%) |
| **Husband age** | 25.6 (5.3) |  | 25.7 (5.3) |  | 26.5 (5.5) |  | 24.9 (5.0) |  | 25.5 (5.4) |  |
| **Wife age** | 17.3 (1.5) |  | 17.4 (1.5) |  | 17.0 (1.6)** |  | 17.4 (1.5) |  | 17.4 (1.5) |  |
| **Husband age at marriage** | 22.4 (5.0) |  | 22.4 (4.7) |  | 23.1 (5.4) |  | 21.9 (4.8) |  | 22.4 (5.0) |  |
| **Husband Education** |  |  |  |  |  |  |  |  |  |  |
| Government school |  | 522 (48.3) |  | 149 (52.8) |  | 136 (52.7)** |  | 126 (45.8) |  | 111 (40.7) |
| Quranic school |  | 224 (20.7) |  | 25 (8.9)** |  | 80 (31.0)** |  | 54 (19.6) |  | 65 (23.8) |
| No school |  | 326 (30.2) |  | 108 (38.3) |  | 40 (15.5) |  | 83 (30.2) |  | 95 (34.8) |
| **Wife Education** |  |  |  |  |  |  |  |  |  |  |
| Government school |  | 382 (35.4) |  | 89 (31.6) |  | 109 (42.3)** |  | 103 (37.5) |  | 84 (30.8) |
| Quranic school |  | 176 (16.3) |  | 7 (2.5)** |  | 84 (32.6)** |  | 31 (11.3) |  | 54 (19.8) |
| No school |  | 514 (47.6) |  | 185 (65.6) |  | 63 (24.4) |  | 137 (49.8) |  | 134 (49.1) |
| **Husband parity** | 1.5 (2.0) |  | 1.6 (1.8) |  | 1.4 (1.8) |  | 1.4 (2.2) |  | 1.43 (2.2) |  |
| **Total household assets (0-6)** | 2.1 (1.2) |  | 1.9 (1.1)* |  | 2.1 (1.2) |  | 2.0 (1.2)* |  | 2.2 (1.1) |  |
| **Husband migration (3+ months/year)** |  | 746 (69.1) |  | 211 (74.8) |  | 146 (54.9) |  | 188 (70.4) |  | 201 (73.6) |
| **Husband Social Norms GEMS score (range: 0-5)** |  |  |  |  |  |  |  |  |  |  |
| T1 | 4.1 (1.1) |  | 3.7 (1.4)** |  | 4.3 (1.0) |  | 4.2 (1.1) |  | 4.2 (0.9) |  |
| T2 | 4.0 (1.1) |  | 4.0 (1.2) |  | 3.7 (1.3)** |  | 4.1 (1.0) |  | 4.2 (0.9) |  |

Abbreviation: SD, standard deviation; GEMS, Gender Equitable Men Scale; T1, baseline data collection; T2, 24-month follow-up data collection
<sup>a</sup>Significance testing conducted to detect baseline differences across study arms using an unadjusted multinomial logistic model regressing each study arm (reference group: control) on baseline characteristics using robust standard errors.
\*p<0.05, \*\*p<0.001

### 4.3. Baseline differences by study retention

Male retention in the study was associated with a variety of baseline factors. Husband study participants were more likely to be retained in the study if they were older (25.9 years among those retained vs. 25.0 years among those not retained at T2, p=0.01), if they had more children with any wife (1.56 children vs. 1.26; p=0.02), and if they did not migrate outside the village for more than three months in the previous 12 months (66.0% migrated vs. 77.6%; p<0.001) [*results not shown*]. Husbands were also more likely to be retained in the study if their wives had received government or Quranic education (34.0% with education vs. 22.4%; (p<0.001) and if the age difference between husband and wife was larger (8.54 years difference vs. 7.72; p=0.01). Study retention did not differ significantly by study arm (Arm 1: 71.0%, Arm 2: 72.9%, Arm 3: 64.4%, Arm 4: 70.3%, p=0.15; data not shown). Factors associated with study retention were used in the creation of propensity weights.

### 4.4. Time trend

The mean SN-GEMS score at T1 was 4.1 [n=1,055; range: 0-5; standard deviation (SD): 1.1] and T2 was 4.0 (n=768; range: 0-5; SD: 1.1), indicating high perceived social acceptability of *inequitable* gender norms. Scores did not significantly differ between T1 and T2 in the entire sample nor among controls alone (mean score among controls at T1: 4.2 vs. at T2: 4.2); no overall temporal trend was detected (unadjusted β: 0.01, 95% confidence intervals (CI): −0.25, 0.28, p=0.92) (Table 2 and 3).

### 4.5. Multilevel Modelling

In both unadjusted and adjusted difference-in-differences models, we found assignment to the small group RMA intervention (Arm 2) to be associated with a significant decrease in perceived social acceptability of inequitable gender norms at follow-up (Table 3). Assignment to RMA small groups among husbands within a given village was associated with a 0.62 lower score on the SN-GEMS (95% CI: −1.05, −0.18) relative to controls, after adjustment for baseline differences. The household visit intervention (Arm 1) was associated with an increase in inequitable gender norms, but this estimate was not significant and imprecise with wide confidence intervals (Table 3). Assignment to the RMA household visits among husbands within a given village was associated with a 0.32 higher score on the SN-GEMS (95% CI: −0.16, 0.80) relative to controls, after adjustment for baseline differences. No significant effects on gender norms were detected for the RMA combination intervention (β: −0.08, 95% CI: −0.42, 0.27).

**Table 3.**
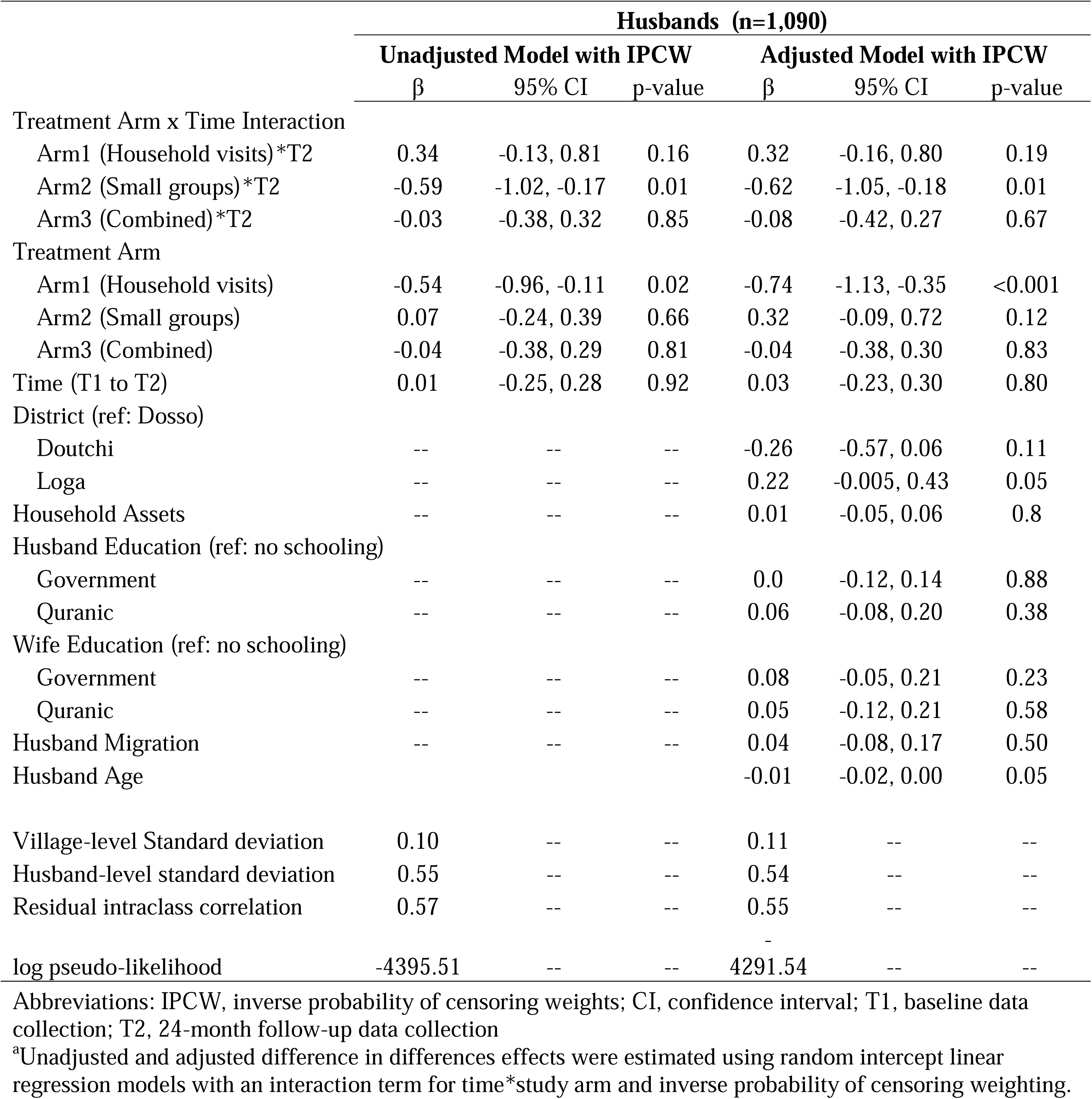
Difference-in-differences effects of the Reaching Married Adolescents Intervention on perceived inequitable gender norms among husbands in Dosso, Niger (2016-2019)^a^.

### 4.6. Sensitivity Analyses Based on Exposure to RMA

We conducted sensitivity analyses to understand RMA effects on SN-GEMS scores when we limited the sample to only those husbands who reported participating in at least one of the intervention components assigned to their district. In adjusted analyses based on exposure to the assigned intervention, we found similar results to those found in the primary analyses: null effects on SN-GEMS for those husbands in Arm 1 exposed to household visits (β: 0.30, 95% CI: −0.20, 0.81), a significant decrease in SN-GEMS scores among husbands in Arm 2 exposed to small groups (β: −0.77, 95% CI: −1.22, −0.32), and null effects among husbands in Arm 3 exposed to at least one intervention component (β: −0.11, 95% CI: −0.51, 0.28) (Table 4). As-treated analyses were not conducted because RMA is a village-level intervention and such analyses would provide bias estimates due to likely spill-over effects. For example, study participants reporting no participation in the assigned intervention component(s) would likely still have had some exposure to the community-level intervention component and spill-over effects from the assigned intervention component being conducted in their district.

**Table 4.**
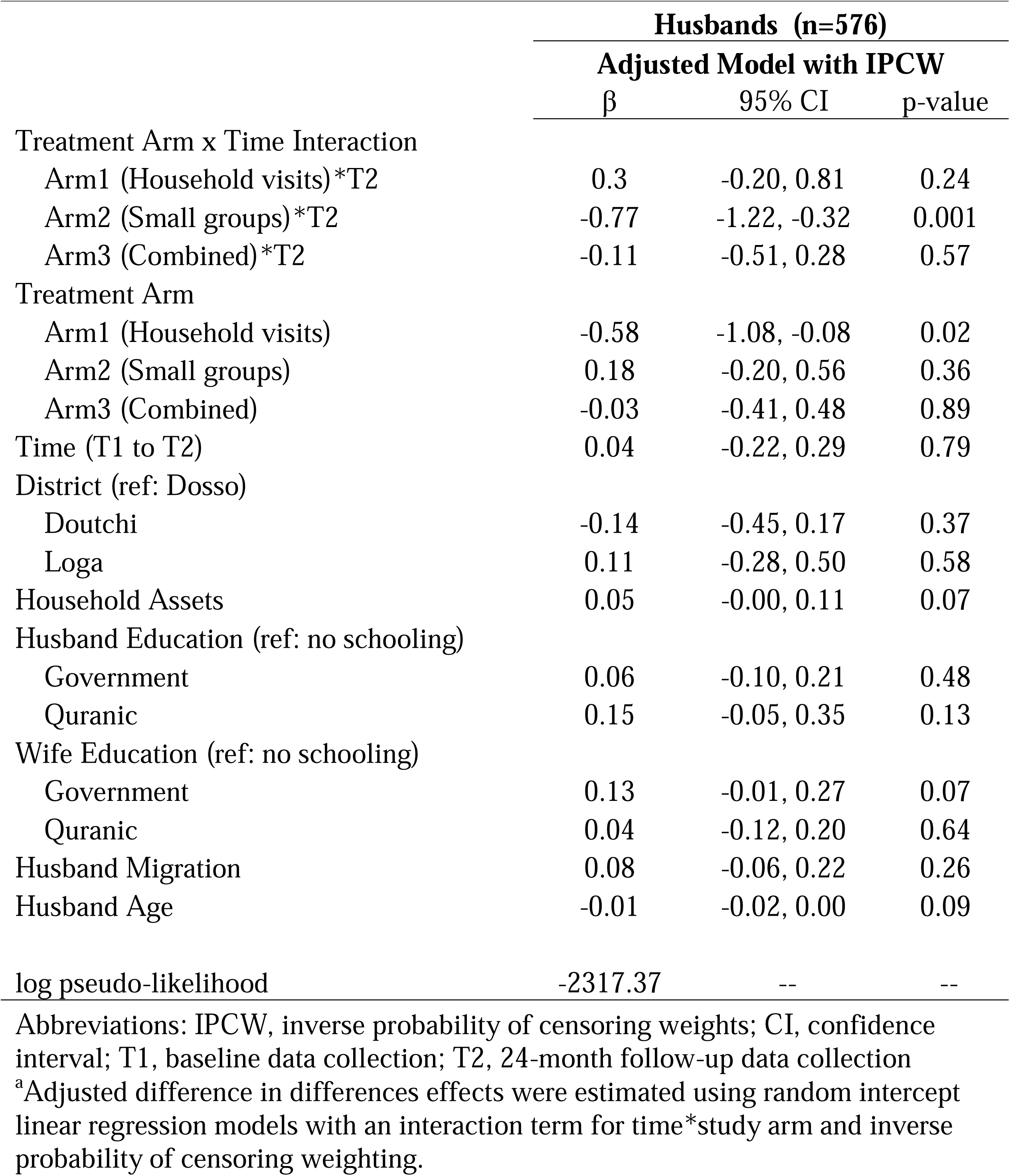
Difference-in-differences effects of the Reaching Married Adolescents Intervention on perceived inequitable gender norms among husbands who participated in the assigned RMA intervention components in Dosso, Niger (2016-2019)^a^.

## 5. DISCUSSION

Results from this cRCT suggest that the small group RMA intervention decreased social norms supporting gender inequitable behaviour among husbands of married adolescent girls in the Dosso region of Niger. Only a limited number of cRCTs and quasi-experimental evaluations of social norm-focused interventions in LMIC contexts have shown direct programmatic effects on social norms (Abramsky et al., 2014; Rowley E, 2020). Even fewer have demonstrated direct effects on both social norms and behavioural outcomes (Abramsky et al., 2014). The present study builds on previous evidence (using the same evaluation data) indicating that the small group RMA intervention reduced IPV among married adolescent girls (Silverman et al., 2022). The present study is the first to demonstrate that the RMA small group intervention also reduced gender inequitable social norms.

Despite the entrenched, deep-seated nature of social norms, the RMA small group intervention approach was successful in creating significant positive shifts in these norms. The small group model, facilitated by local CHWs, aimed to provide education and a space for men to gather, discuss, and possibly challenge one another’s thinking around gender roles. The resulting shifts in norms observed in this study provide empirical evidence to support current theoretical understanding of how social norms change; social norms are socially constructed concepts of acceptable behaviour within a community and are most amenable to change in social contexts. Other interventions that have been shown to change social norms or aggregated individual attitudes, like Sasa!, also utilize male discussion groups to shift social norms, but typically also engage the wider community through mobilizing efforts (Abramsky et al., 2014; Cislaghi & Heise, 2018). The RMA intervention offers a simple approach to social norms change that is feasible to implement in one of the most resource-limited settings among LMICs.

Social norms interventions have been shown to be effective when they provide information or correct misinformation about peer’s behaviours or provide space for renegotiation of social norms within a social group (Miller & Prentice, 2016). It aligns, then, that receipt of RMA household visits, an intervention delivered to each individual of a married couple without information about peer behaviour or opportunity for the exchange of ideas among peers, did not show evidence of changing perceived inequitable gender norms. The RMA intervention, including household visits, was designed with the aim of increasing contraceptive use (i.e., social norms were not the primary outcome) and evidence indicates household visits were effective at doing so (Silverman et al., 2022). The non-significant effect on perceived inequitable gender norms associated with household visits, though, suggests that these visits provided little opportunity for husbands to see change within their peers around issues of gender equity. Similar null effects were observed in the combination arm. As the combination arm offered two intervention components (household visits and small groups) with opposing effects on social norms, these effects likely counter-balanced one another, resulting in no effect on social norms in this arm.

This study is not without limitations. First, despite treatment randomization, baseline differences in treatment and control arms exist because the first level of randomization was linked to district. This design decision was due to real-world constraints of randomization, which, in this case, came from the Ministry of Health’s requirement that randomization be conducted at the district level. Baseline differences likely resulted from important social, economic, and cultural differences among district residence. To address this, control villages were randomly selected from all of the three intervention districts such that each district is represented by a third of the control villages. We also utilized a difference-in-difference analytical approach and controlled for district and other baseline differences, mostly related to randomization at the district level. Still, unmeasured confounding may obscure these results. For example, at baseline, Arm 1 had significantly lower levels of Quranic schooling relative to controls. Importantly, this arm also had significantly lower scores on the SN-GEMS at baseline relative to controls, suggesting lower acceptance of inequitable gender norms relative to other arms, which could be related to the lower levels of Quranic education in these villages, and may contribute to the imprecise estimation of effects on social norms for Arm 1. More scrutiny of these and other factors that may have contributed to unmeasured confounding is needed to confirm results on RMA household visits.

Second, there was notable loss to follow-up among husband participants at T2, which could have led to selection bias. To minimize the impact of this source of selection bias, we utilized IPCW in our models to re-weight the data so that it more closely approximates the data had those participants not been lost. Third, interpretation of random intercept models is the change in score between individuals receiving treatment and those receiving control *within a cluster*. In this study design, all residents in a village received either treatment or control, never a mix of the two. Therefore, the random effects model interpretations reflect a theoretical within village effect that was not possible in reality. Finally, these results only represent the effects of the RMA intervention on inequitable gender norms in a probability sample of husbands of married adolescent girls in the rural Dosso region of Niger. While these results are not representative of other regions in sub-Saharan Africa, they do suggest that if positive impact can be achieved in this setting, the small groups elements of the RMA intervention may be promising in other similarly resourced, culturally-related settings. Future research in other settings is needed to assess if effects can be replicated.

## 6. CONCLUSIONS

These results provide preliminary evidence that the RMA small group intervention approach is effective at reducing inequitable gender norms, one of very few interventions in LMIC demonstrating such effects. While further careful investigation of unmeasured confounding is needed, these results also suggest that the household visit and combination RMA intervention did not change perceptions of inequitable gender norms, possibly due to the lack of facilitated social exchange in household visits. Inequitable gender norms are understood to be an important, though difficult to change, upstream risk factor for a variety of adverse health outcomes, including intimate partner violence, reproductive coercion, child marriage, and maternal mortality. The potential ripple effect of an intervention that decreases inequitable gender norms among this high priority population of husbands of married adolescent girls in Niger could provide long term benefit across multiple related health outcomes. Moreover, as a low-cost, simple, scalable, and transferrable intervention with evidence of being able to change such gender norms, this CHW-based small group intervention approach could be extremely valuable to the field of public health for reducing the negative impact of inequitable gender norms on health and wellbeing in similar settings.

## ETHICAL STATEMENT

### Ethics approval and consent to participate

Our study was approved by both the University of California San Diego School of Medicine Institutional Review Board (Project 160407S) and the Research Ethics Board of the Niger Ministry of Health (Project 011/2016/CCNE). Consent was obtained from all participants who provided data.

### Consent for publication

Not applicable

### Data availability

The data that support the findings of this study are available from the corresponding author, SB, upon reasonable request.

### Author contributions

**Sabrina C. Boyce:** conceptualization, methodology, formal analysis, investigation, writing – original draft, editing, visualization, project administration, funding acquisition; **Alexandra M. Minnis:** conceptualization, writing-review & editing, supervision; **Julianna Deardorff:** conceptualization, writing-review & editing, supervision; **Sandra McCoy:** conceptualization, writing-review & editing, supervision; **Sneha Challa:** investigation, writing – review & editing, project administration; **Nicole E. Johns:** data curation, formal analyses, writing – review & editing; **Sani Aliou:** investigation, resources, project administration, writing – review & editing; **Mohamad Brooks:** investigation, project administration, writing – review & editing; **Abdoul-Moumouni Nouhou:** investigation, resources, writing – review & editing, project administration; **Holly Baker:** conceptualization, methodology, writing – review & editing; **Jay G. Silverman:** conceptualization, methodology, project administration, writing – review & editing, supervision, funding acquisition.

